# Point-of-care testing for COVID-19 using SHERLOCK diagnostics

**DOI:** 10.1101/2020.05.04.20091231

**Authors:** Julia Joung, Alim Ladha, Makoto Saito, Michael Segel, Robert Bruneau, Mee-li W. Huang, Nam-Gyun Kim, Xu Yu, Jonathan Li, Bruce D. Walker, Alexander L. Greninger, Keith R. Jerome, Jonathan S. Gootenberg, Omar O. Abudayyeh, Feng Zhang

## Abstract

The recent outbreak of the novel coronavirus SARS-CoV-2, which causes COVID-19, can be diagnosed using RT-qPCR, but inadequate access to reagents and equipment has slowed disease detection and impeded efforts to mitigate viral spread. Alternative approaches based on combinations of isothermal amplification and CRISPR-mediated detection, such as the SHERLOCK (<u>S</u>pecific <u>H</u>igh <u>S</u>ensitivity <u>E</u>nzymatic <u>R</u>eporter Un<u>LOCK</u>ing) technique, offer reduced dependence on RT-qPCR equipment, but previously reported methods required multiple fluid handling steps, complicating their deployment outside clinical labs. Here we developed a simple test chemistry called STOP (SHERLOCK Testing in One Pot) for detecting SARS-CoV-2 in one hour that is suitable for point-of-care use. This simplified test, STOPCovid, provides sensitivity comparable to RT-qPCR-based SARS-CoV-2 tests and has a limit of detection of 100 copies of viral genome input in saliva or nasopharyngeal swabs per reaction. Using lateral flow readout, the test returns result in 70 minutes, and using fluorescence readout, the test returns result in 40 minutes. Moreover, we validated STOPCovid using nasopharyngeal swabs from COVID-19 patients and were able to correctly diagnose 12 positive and 5 negative patients out of 3 replicates. We envision that implementation of STOPCovid will significantly aid “test-trace-isolate” efforts, especially in low-resource settings, which will be critical for long-term public health safety and effective reopening of the society.

## Introduction

Rapid point-of-care (POC) tests capable of operating in any low-resource setting, including at home, are needed to adequately combat the COVID-19 pandemic and reopen society. POC tests for SARS-CoV-2, the novel coronavirus causing COVID-19 disease, that have been [authorized by the U.S. Food and Drug Administration (FDA)](https://www.fda.gov/medical-devices/emergency-situations-medical-devices/emergency-use-authorizations#covid19ivd), including the Abbott ID NOW and Cepheid GeneXpert, require specialized and expensive instrumentation, high level technical expertise, are expensive, and prohibit widespread use. Therefore scalable, affordable, and easy-to-use POC tests remain an urgent need for the fight again COVID-19.

Previously, we developed a sensitive and specific nucleic detection technology called SHERLOCK (<u>S</u>pecific <u>H</u>igh <u>S</u>ensitivity <u>E</u>nzymatic <u>R</u>eporter Un<u>LOCK</u>ing) (Gootenberg et al., 2017, 2018), which achieves detection of DNA or RNA virus signatures through two consecutive reactions: (1) amplification of the viral RNA using an isothermal amplification reaction, and (2) detection of the resulting amplicon using CRISPR-mediated collateral reporter unlocking. During the early period of the COVID-19 outbreak, we developed a SHERLOCK-based test for SARS-CoV-2 ([SHERLOCK-Covid](https://www.broadinstitute.org/files/publications/special/COVID-19%20detection%20(updated).pdf)). Additional CRISPR-based tests have also been recently developed (Broughton et al., 2020; Ding et al., 2020; Guo et al., 2020; Lucia et al., 2020), but similar to the SHERLOCK-Covid test, all involve two separate reaction steps, liquid handling, and opening of tubes. These steps not only add complexity but also significantly increase the likelihood of sample cross-contamination, prohibiting their use outside well-controlled laboratory environments.

Other isothermal pre-amplification methods, such as Loop-mediated Isothermal Amplification (LAMP), are also being developed as POC tests (Zhang et al., 2020), but these rely on amplification that can be nonspecific. Unlike isothermal amplification-only tests, SHERLOCK provides two rounds of sequence-specific detection, first through primer-specific isothermal amplification, and then via guide RNA-mediated, sequence-specific detection of the viral sequence.

To make SHERLOCK ready for use in POC contexts, we developed a simple testing chemistry called STOP (SHERLOCK Testing in One Pot), which does not require sample extraction and can be performed at a single temperature with one fluid handling step and a simple visual readout similar to a pregnancy test (Figure 1A). Here we report a STOP test for COVID-19 called STOPCovid.

**Figure 1:**
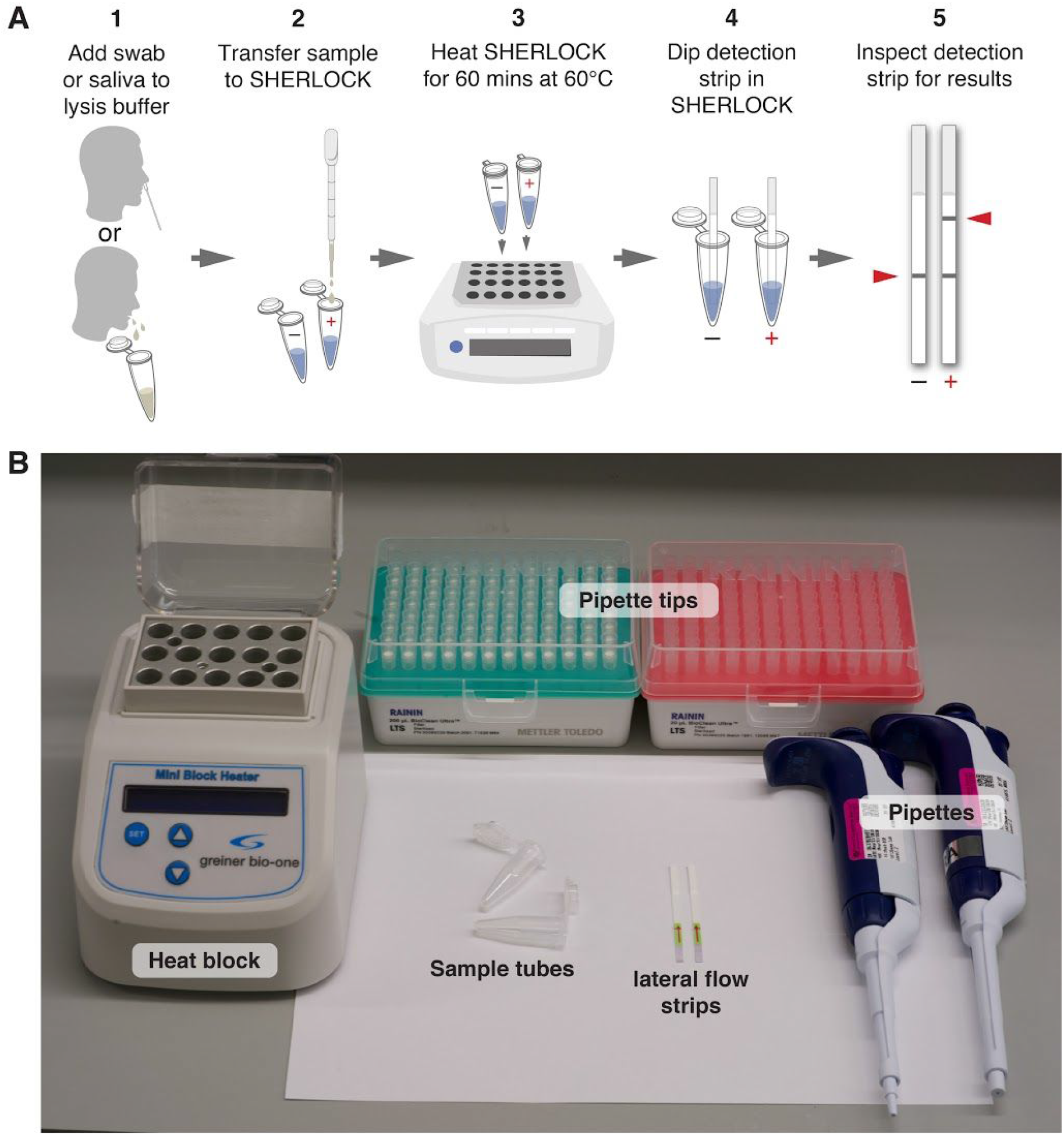
Overview of the STOPCovid method. A. Schematic of the one-step STOPCovid test. A nasopharyngeal swab or saliva is transferred into the lysis buffer. Sample is transferred to negative control and test SHERLOCK reactions and heated for 60 mins at 60°C. Detection strips are then dipped into each reaction for 2 minutes for lateral flow readout. B. Equipment and consumables needed for running STOPCovid, including a heat block, pipettes, pipette tips, sample tubes, and lateral flow strips.

The STOPCovid test works in the following three steps, without requiring separate viral RNA extraction and only simple laboratory equipment (Figure 1B).

|  |  |  |
| --- | --- | --- |
| *Step (1)* | 10 mins at 22 °C;<br>10 mins at 60 °C;<br>Or 5 mins at 95 °C | lysis of virus-containing patient sample using QuickExtract to release viral RNA; |
| *Step (2)* | 1 hr at 60 °C | detection of viral RNA using STOPCovid reaction; |
| *Step (3)* | 2 mins at 22 °C | visual read out of the detection result by eye using a commercially-available paper dipstick. |

## Results

In order to turn the classically two-step SHERLOCK into a single-step reaction by integrating the isothermal amplification step with the CRISPR-mediated detection step, we sought to establish a common reaction chemistry capable of supporting both steps. Due to the supply chain constraints for the commercially-available recombinase polymerase amplification (RPA) reagents and difficulties in producing a rapid one-pot RPA test for sensitive RNA detection, we chose loop-mediated isothermal amplification (LAMP) reaction for amplifying the viral RNA.

The requisite enzymes for LAMP are more readily available from a number of commercial suppliers and the LAMP buffers are defined and amenable to systematic optimization with Cas enzymes.

To determine the optimal combination of LAMP primers and guides, we designed 29 sets of LAMP primers targeting different regions of the SARS-CoV-2 genome and identified the best primer set for amplifying gene N, which encodes the nucleoprotein (Figure 2A). As LAMP operates at a higher temperature than RPA (55-65 °C compared to 37-42 °C), a one-pot reaction demands a Cas enzyme with collateral activity that is thermostable. *Alicyclobacillus acidoterrestris* Cas12b (AacCas12b), which has been shown to be thermostable at 55C (Shmakov et al., 2015), has been incorporated into a one-pot detection assay with LAMP (Li et al., 2019), but because it is unstable at the high temperature required for LAMP, we did not find adequate sensitivity and performance. Of the various Cas proteins we explored, Cas12b from *Alicyclobacillus acidiphilus* (AapCas12b) (Teng et al., 2018) maintained sufficient activity in the same temperature range as LAMP (Figure 2B).

**Figure 2:**
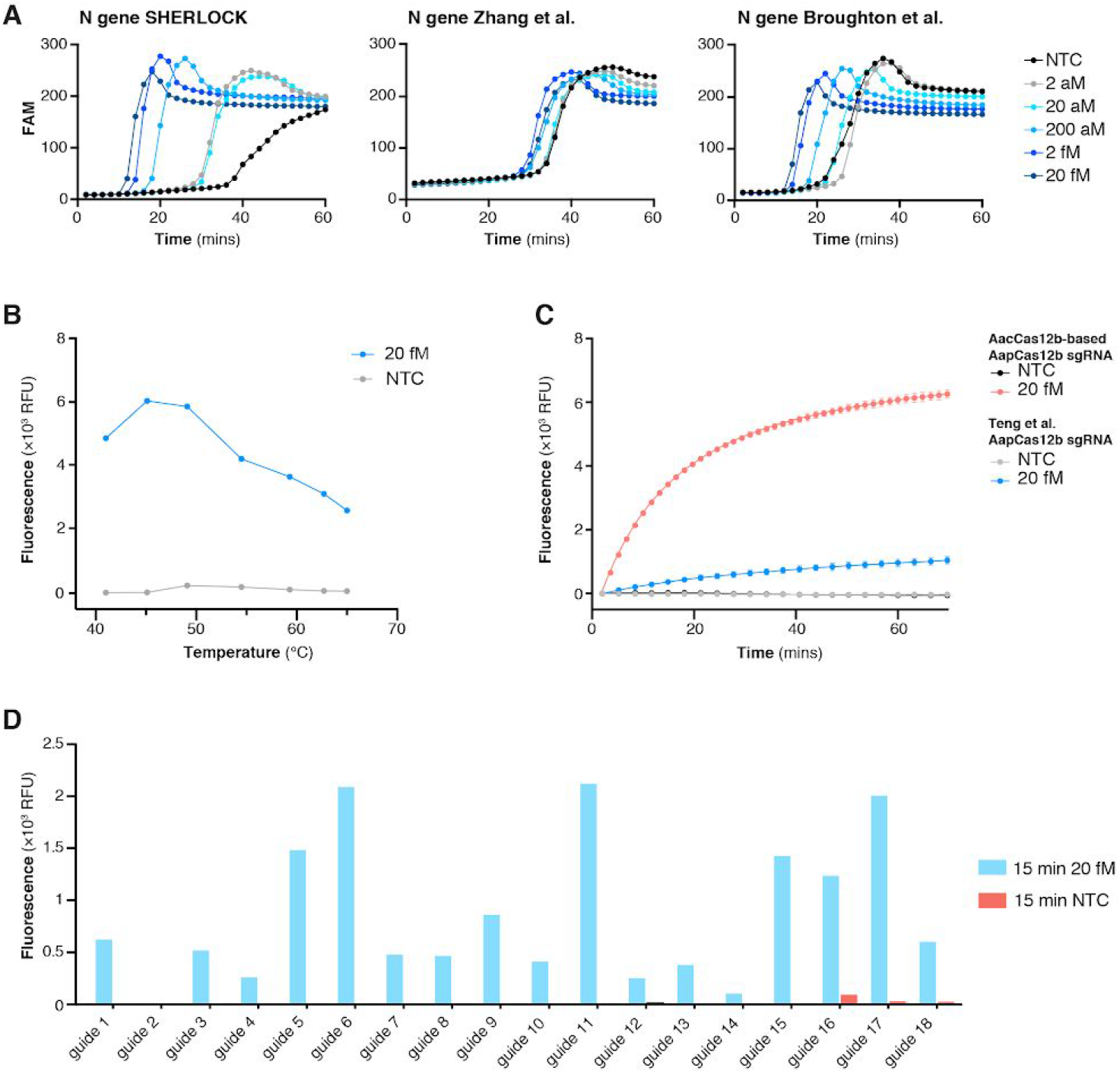
Development of STOPCovid using RT-LAMP and thermophilic AapCas12b. A. Comparison of the STOPCovid N gene LAMP primer set to two established LAMP primer sets measured by real-time fluorescence across a range of concentrations of SARS-CoV-2 genomic standards. NTC, no template control. B. Temperature dependence of AapCas12b collateral activity when incubated with RT-LAMP amplified 20 fM SARS-CoV-2 genomic standard (blue) or no template control, NTC (gray). Collateral activity was measured as 5-minute end point fluorescence after incubation. C. Comparison of AapCas12b collateral activity with either a previously published sgRNA scaffold (Teng et al., 2019) (blue) or an AacCas12b-based scaffold (red) when incubated with RT-LAMP amplified 20 fM SARS-CoV-2 genomic standard. D. Collateral activity of AapCas12b using different guides when incubated with RT-LAMP amplified 20 fM SARS-CoV-2 genomic standard.

However, because the AapCas12b locus did not contain a CRISPR array, the published single guide RNA (sgRNA) for AapCas12b (Teng et al., 2018) used a direct repeat (DR) sequence from *Alicyclobacillus macrosporangiidus* Cas12b, which could impede activity. To remedy this, we searched for alternative orthologs with similar protein sequences to AapCas12b and found that AacCas12b shared a 95% sequence homology (Shmakov et al., 2015). Additionally, the AacCas12b tracrRNA and predicted AapCas12b tracrRNA are 97% identical. Given the high degree of similarity between AapCas12b and AacCas12b protein and tracrRNA, we surmised that the sgRNA for AacCas12b should closely match the cognate AapCas12b DR-tracrRNA hybrid. Indeed, reactions combining AapCas12b enzyme with AacCas12b scaffold sgRNA produced more robust and specific nuclease activity compared to the previously published AapCas12b sgRNA (Figure 2C).

For the best LAMP amplicon, we systematically evaluated 18 sgRNAs to identify the optimal combination of primers and guide sequence (Figure 2D). Using this combination in a one-pot reaction, we found that AapCas12b generated faster and higher collateral activity than AacCas12b protein (Figure 3A). We further optimized one-pot reaction components by screening 94 additives to improve thermal stability of the one pot reaction (Figure 3B), finding that addition of taurine significantly improved reaction kinetics (Figure 3C).

**Figure 3:**
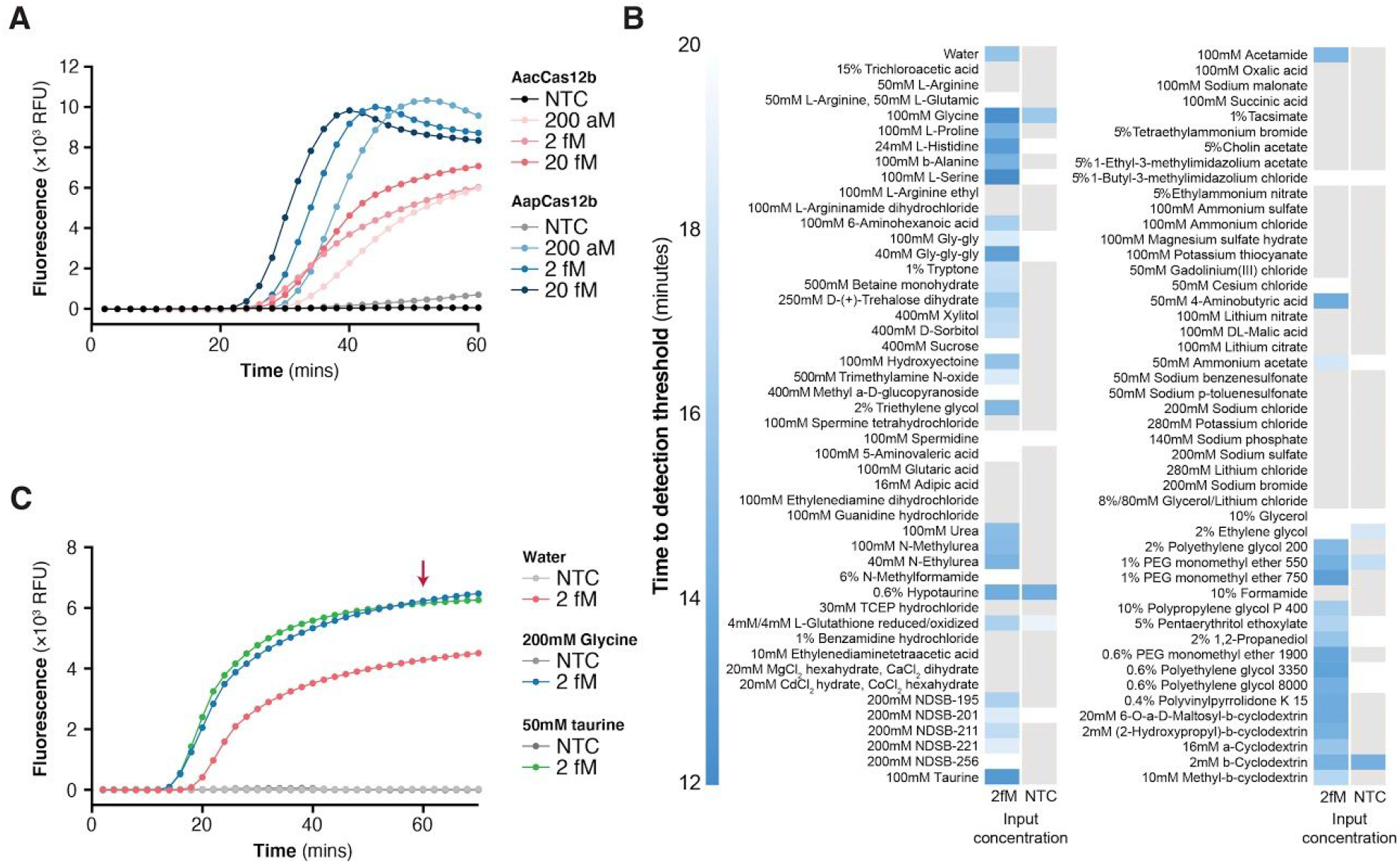
Optimization of STOPCovid with additive reagents. A. Comparison of STOPCovid performance (monitored by real-time fluorescence) when using either AacCas12b (red) or AapCas12b (blue) and varying amounts of SARS-CoV-2 genomic standards. AacCa12b sgRNA was used for both enzymes. NTC, no template control. B. Heat map comparing a panel of chemical additives for effects on the performance of STOPCovid for detection of 2 fM SARS-CoV-2 genomic standard. C. Comparison of STOPCovid performance (monitored by real-time fluorescence) with either glycine or taurine additives at 2 fM SARS-CoV-2 genomic standard.

We profiled the optimized reaction with a lateral flow readout and an RNA extraction-free input using SARS-CoV-2 genome standards spiked into pooled healthy saliva or nasopharyngeal (NP) swabs to determine the limit of detection (LOD), ideal incubation temperature, readout time, and robustness. We found that the LOD of the reaction was 100 copies of SARS-CoV-2 (Figure 4A,B) for readout using one strip. This LOD was reliable and reproducible over 30 replicates (Figure 4A,B). The ideal incubation parameters were 60°C for at least 50 minutes for lateral flow (Figure 4C,D), and the reaction components could be formulated as a mastermix and maintained functionality after 6 freeze-thaw cycles (Figure 4E). The assay exhibited no cross-reactivity with the SARS-CoV or MERS-CoV genomes (Figure 4F) and could be performed using either a standard heat block or via a water bath maintained by a commercially-available low-cost (under $40USD) sous-vide cooker (Figure 4G,H).

**Figure 4:**
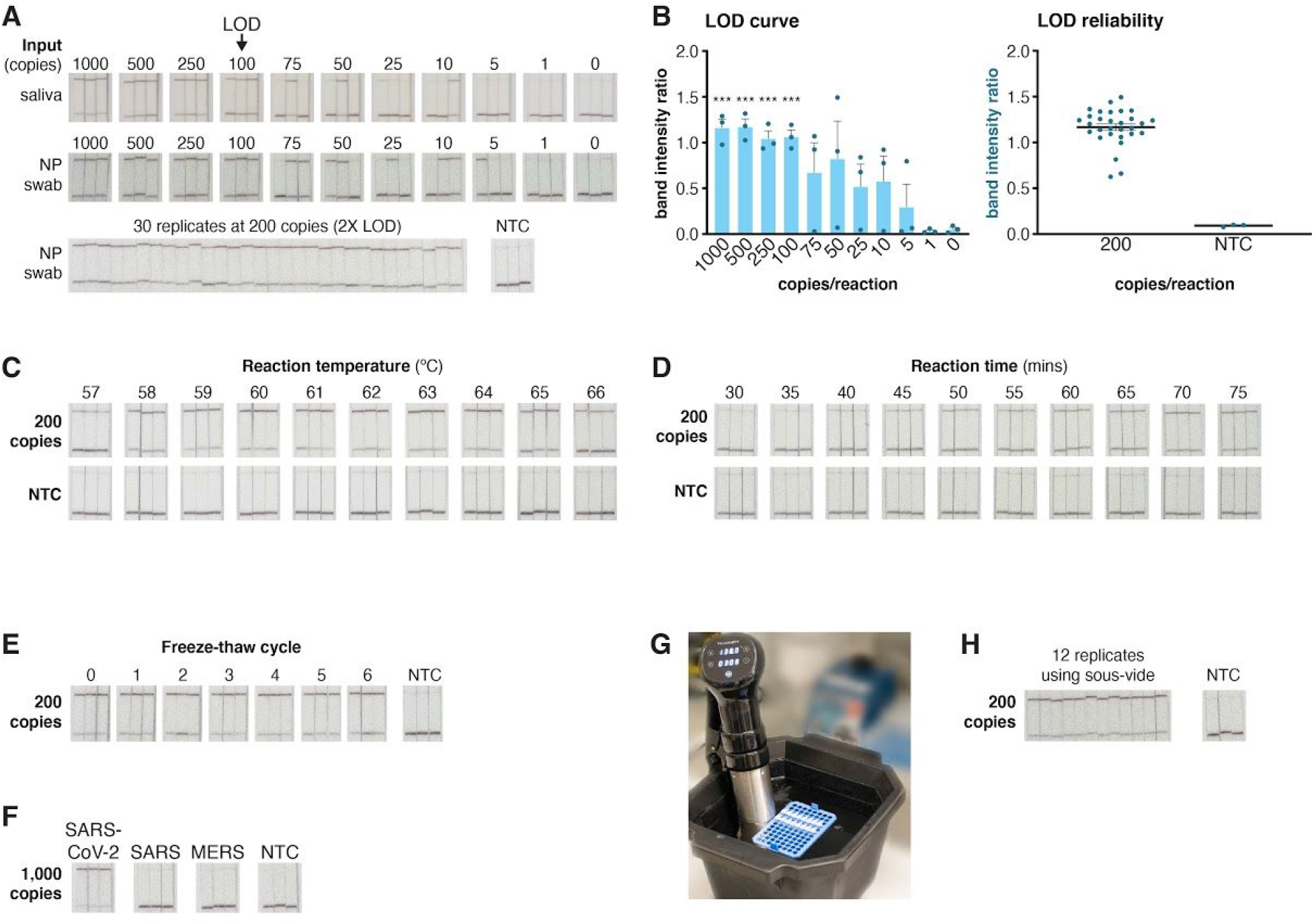
STOPCovid performance on lateral flow strips. A. Determination of the limit of detection (LOD) for STOPCovid with lateral flow readout using three replicates per condition (top). Synthetic genomic standards spiked into saliva or nasopharyngeal (NP) swab was used for input. All three replicates could be positively detected (top band on strip) down to 100 SARS-CoV-2 standard genomes per reaction. At twice the determined LOD of 100 genomic copies per reaction in NP swab, STOPCovid yielded positive results for 30 replicates (bottom). NTC, no template control. B. Quantification of the band intensity ratio (top band/bottom band) for the LOD determination reactions (left) and 30x replicates (right) from panel A. ***, p < 0.001. C. Effect of reaction temperature on STOPCovid lateral flow detection for 200 SARS-CoV-2 copies per reaction. D. Effect of reaction incubation time on STOPCovid lateral flow detection for 200 SARS-CoV-2 copies per reaction. E. Effect of mastermix freeze-thaw cycles on STOPCovid lateral flow detection for 200 SARS-CoV-2 copies per reaction. F. Evaluation of cross-reactivity for COVID-19 STOPCovid lateral flow test for SARS and MERS N genes. All inputs were at 1,000 copies per reaction. G. Example STOPCovid setup using a 60°C water bath powered by a simple commercial sous-vide device. H. STOPCovid lateral flow detection of SARS-CoV-2 at 200 copies per reaction using a 60°C sous-vide water bath.

We evaluated STOPCovid detection using viral transport media from NP swabs obtained from 12 SARS-CoV-2 RNA positive and 5 negative patients (Figure 5A-C). Our assay reliably detected SARS-CoV-2 in patient samples: 3 out of 3 replicates for 11/12 patient samples, and at least 2 out of 3 replicates for 12/12 patient samples. STOPCovid also reliably returned negative results for all 5 negative patient samples (0 out of 3 replicates for each sample).

**Figure 5:**
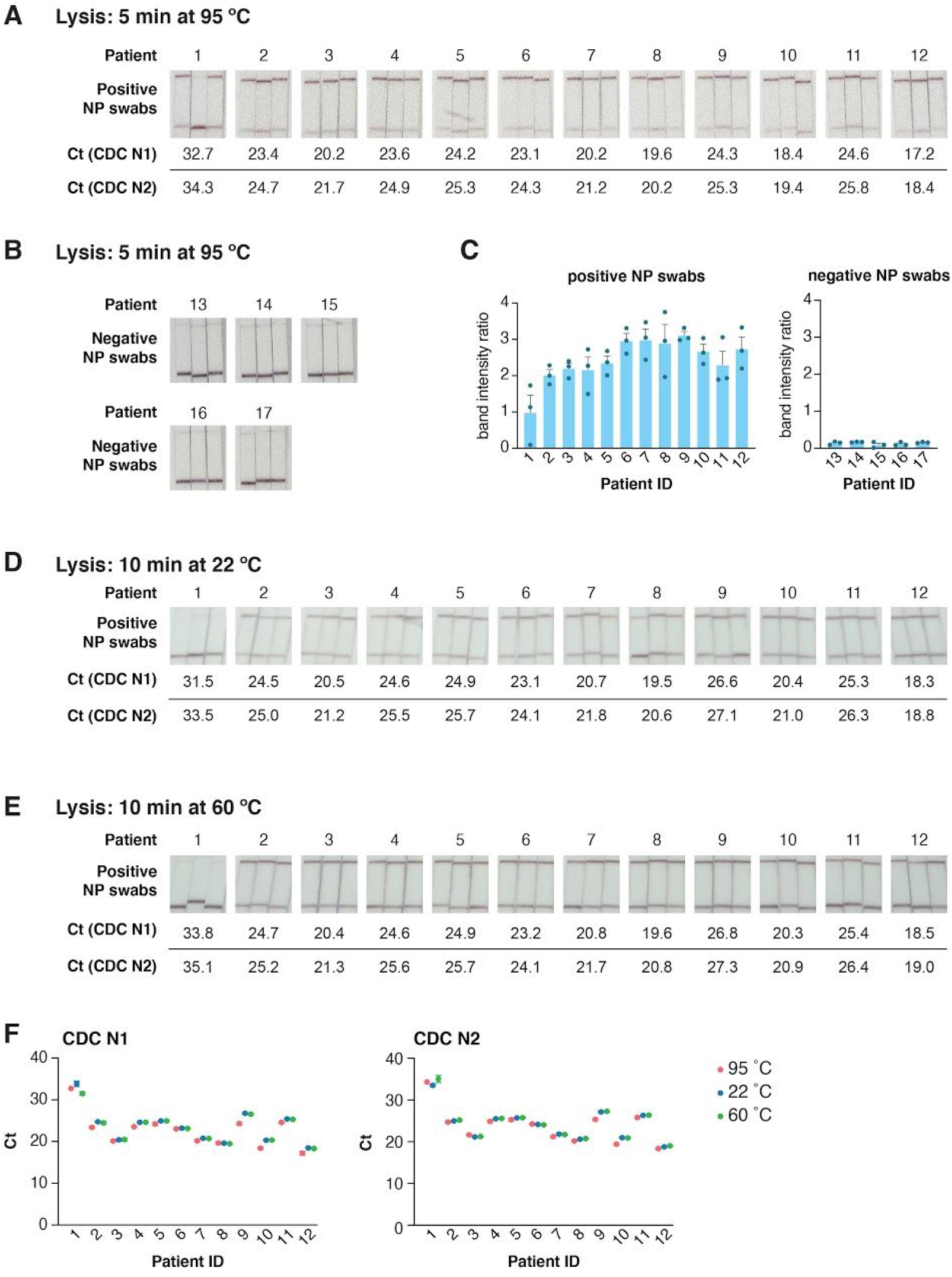
STOPCovid with SARS-CoV-2 patient nasopharyngeal swab samples. A. STOPCovid results for 12 unique SARS-CoV-2 positive patient nasopharyngeal swab samples in triplicate. Prior to STOPCovid, 30 μL of VTM elution from nasopharyngeal swabs was lysed by adding an equal volume of QuickExtract and heating for 5 minutes at 95°C. Ct values determined by RT-qPCR using the CDC N1 (top) and N2 (bottom) assays for each patient sample are shown below. B. Same as panel A, for and 5 SARS-CoV-2 negative patient nasopharyngeal swab samples. C. Quantification of the band intensity ratios of lateral flow results from panels A and B. D-E. Alternative lysis methods on STOPCovid results on the same set of 12 positive patients as panel A in triplicates. Prior to STOPCovid, 30 μL of VTM elution from nasopharyngeal swabs were lysed in QuickExtract for 10 minutes at 22°C (D) or 60°C (E). Ct values determined by RT-qPCR using the CDC N1 (top) and N2 (bottom) assays for each patient sample are shown below. F. Comparison of Ct values for samples lysed at 95°C, 22°C and 60°C from panels A, D, and E. For patients 9 and 10, due to the low volume of samples available, samples lysed at 22°C and 60°C were diluted 1:2 prior to STOPCovid and RT-qPCR.

To further simplify the assay workflow, we tested if lysis with QuickExtract at room temperature (22 °C) or at one-pot incubation temperature (60 °C) for 10 mins would be sufficient for detection. As QuickExtract contains Proteinase K that inhibits SHERLOCK without heat inactivation at 95 °C, we added Proteinase K Inhibitor to the SHERLOCK and RT-qPCR reactions. In both cases, we could identify 11/12 positive patient samples (Figure 5D,E). Comparison of RT-qPCR Ct values between the lysis methods suggested that both lysis methods are viable alternatives for streamlining the assay workflow, though with a slight decrease (0.2/0.4 Ct at 60°C and 0.4/0.7 Ct for 22°C for CDC N1/N2 probe sets) in sensitivity (Figure 5F).

## Materials and Methods

### Design of LAMP and SHERLOCK reactions

We designed LAMP amplification primers and SHERLOCK AapCas12b guide RNAs to target the N gene of SARS-CoV-2. The N gene is known to be present at higher copy numbers than other segments of the SARS-CoV-2 genome, which helps to increase the detection sensitivity (Kim et al., 2020). Below are the LAMP primer sequences and SHERLOCK AapCas12b guide RNAs:

LAMP Primers
F3: 5′– GCTGCTGAGGCTTCTAAG –3′
B3: 5′– GCGTCAATATGCTTATTCAGC –3′
FIP: 5′– GCGGCCAATGTTTGTAATCAGTAGACGTGGTCCAGAACAA –3′
BIP: 5′– TCAGCGTTCTTCGGAATGTCGCTGTGTAGGTCAACCACG –3′
Loop Forward: 5′– CCTTGTCTGATTAGTTCCTGGT –3′
Loop Reverse: 5′– TGGCATGGAAGTCACACC –3′

> 5′–UCUAGAGGACAGAAUUUUUCAACGGGUGUGCCAAUGGCCACUUUCCAGGUGG CAAAGCCCGUUGAGCUUCUCAAAUCUGAGAAGUGGCAC**<u>CGAAGAACGCUGAAGCGCUG</u>** -3' (The spacer targeting N gene is underlined.)

> MAVKSMKVKLRLDNMPEIRAGLWKLHTEVNAGVRYYTEWLSLLRQENLYRRSPNGDGEQ ECYKTAEECKAELLERLRARQVENGHCGPAGSDDELLQLARQLYELLVPQAIGAKGDAQ QIARKFLSPLADKDAVGGLGIAKAGNKPRWVRMREAGEPGWEEEKAKAEARKSTDRTAD VLRALADFGLKPLMRVYTDSDMSSVQWKPLRKGQAVRTWDRDMFQQAIERMMSWESWNQ RVGEAYAKLVEQKSRFEQKNFVGQEHLVQLVNQLQQDMKEASHGLESKEQTAHYLTGRA LRGSDKVFEKWEKLDPDAPFDLYDTEIKNVQRRNTRRFGSHDLFAKLAEPKYQALWRED ASFLTRYAVYNSIVRKLNHAKMFATFTLPDATAHPIWTRFDKLGGNLHQYTFLFNEFGE GRHAIRFQKLLTVEDGVAKEVDDVTVPISMSAQLDDLLPRDPHELVALYFQDYGAEQHL AGEFGGAKIQYRRDQLNHLHARRGARDVYLNLSVRVQSQSEARGERRPPYAAVFRLVGD NHRAFVHFDKLSDYLAEHPDDGKLGSEGLLSGLRVMSVDLGLRTSASISVFRVARKDEL KPNSEGRVPFCFPIEGNENLVAVHERSQLLKLPGETESKDLRAIREERQRTLRQLRTQL AYLRLLVRCGSEDVGRRERSWAKLIEQPMDANQMTPDWREAFEDELQKLKSLYGICGDR EWTEAVYESVRRVWRHMGKQVRDWRKDVRSGERPKIRGYQKDVVGGNSIEQIEYLERQY KFLKSWSFFGKVSGQVIRAEKGSRFAITLREHIDHAKEDRLKKLADRIIMEALGYVYAL DDERGKGKWVAKYPPCQLILLEELSEYQFNNDRPPSENNQLMQWSHRGVFQELLNQAQV HDLLVGTMYAAFSSRFDARTGAPGIRCRRVPARCAREQNPEPFPWWLNKFVAEHKLDGC PLRADDLIPTGEGEFFVSPFSAEEGDFHQIHADLNAAQNLQRRLWSDFDISQIRLRCDW GEVDGEPVLIPRTTGKRTADSYGNKVFYTKTGVTYYERERGKKRRKVFAQEELSEEEAE LLVEADEAREKSVVLMRDPSGIINRGDWTRQKEFWSMVNQRIEGYLVKQIRSRVRLQES ACENTGDI*

### Specimen and nucleic acid extraction

Two types of patient samples have been tested for compatibility with STOPCovid. All samples should be collected and processed according to the appropriate biosafety procedure.

a. *RNA extracted from patient samples:* Please reference the [2020 CDC COVID-19 test protocol](https://www.cdc.gov/coronavirus/2019-ncov/lab/rt-pcr-detection-instructions.html) for details on specimen collection and subsequent nucleic acidextraction. The input for this protocol can be the same extracted nucleic acid as used in RT-qPCR assays. If starting with extracted RNA, begin with Step (2).
b. *Saliva or nasopharyngeal (NP) swabs:* Saliva or NP swabs dipped in viral transport media (VTM) or VTM equivalents can be directly used.

### Reagents

For Step (1) - lysis of viral sample:

- QuickExtract DNA Extraction Solution ([QE09050](https://www.lucigen.com/QuickExtract-DNA-Extraction-Solution/)), Lucigen. Once thawed, aliquot and store at −20 °**C** to avoid >3 freeze-thaw cycles.

For Step (2) - STOPCovid detection reaction:

- Bst 2.0 WarmStart® DNA Polymerase ([M0538L](https://www.neb.com/products/m0538-bst-20-warmstart-dna-polymerase)), New England BioLabs
- WarmStart® RTx Reverse Transcriptase ([M0380L](https://www.neb.com/products/m0380-warmstart-rtx-reverse-transcriptase#Product%20Information)), New England BioLabs
- 10X Isothermal Amplification Buffer ([B0374S](https://www.neb.com/products/b0374-isothermal-amplification-buffer-ii#Product%20Information)), New England BioLabs, supplied with M0538L and M0380L
- 100 mM MgSO_4_ ([B1003S](https://www.neb.com/products/b1003-magnesium-sulfate-mgso4-solution#Product%20Information)), New England BioLabs, supplied with M0538L and M0380L
- 10 mM Deoxynucleotide (dNTP) Solution Mix ([N0447L](https://www.neb.com/products/n0447-deoxynucleotide-dntp-solution-mix#Product%20Information)), New England BioLabs
- Taurine ([86329-100G](https://www.sigmaaldrich.com/catalog/product/sigma/86329)), Millipore Sigma
- AapCas12b protein purified according to [Kellner et al.. Nature Protocols 2019](https://www.nature.com/articles/s41596-019-0210-2). stored as 10 μL aliquots at 2 mg/mL.
- Guide RNA for detecting N gene can be ordered from [Synthego](https://orders.synthego.com/products/crisprevolution-custom-rna-9/#/tubes)
- Reporter DNA for lateral flow read out (Lateral Flow Reporter: /56-FAM/TTTTTTT/3Bio/), can be ordered from [IDT](https://www.idtdna.com/site/order/oligoentry)
- (Optional) Proteinase K Inhibitor ([539470-10MG](https://www.emdmillipore.com/US/en/product/Proteinase-K-Inhibitor-Calbiochem,EMD_BIO-539470)), Millipore Sigma. Resuspend 10 mg of Proteinase K Inhibitor with 150 μL of DMSO to make the stock solution. Dilute stock solution 1:100 with ddH_2_O to make working aliquots. Store both stock and working solutions at −20 °**C**
- (Optional) Reporter DNA for fluorescent readout (Fluorescent Reporter: /5HEX/TTTTTTT/3IABkFQ/), can be ordered from [IDT](https://www.idtdna.com/site/order/oligoentry))
- 10X LAMP Primer Mix can be prepared as follows:

| LAMP Primer (100 µM, in ddH<sub>2</sub>O) | Amount (µL) |
| --- | --- |
| F3 | 2 |
| B3 | 2 |
| FIP | 16 |
| BIP | 16 |
| Loop F | 4 |
| Loop B | 4 |
| ddH<sub>2</sub>O | 56 |
| **Total** | **100** |

A Sherlock mastermix can be prepared as follows:

|  | **Initial**<br>**concentration** | **Final**<br>**concentration** | **Amount**<br>**(uL)** |
| --- | --- | --- | --- |
| Isothermal<br>amplification<br>buffer | 10 X | 1 X | 5 |
| dNTPs | 10 mM | 1.4 mM | 7 |
| MgSO<sub>4</sub> | 100 mM | 8 mM | 4 |
| WS Bst 2.0 | 8000 units/mL | 320 units/mL | 2 |
| WS RTx | 15,000<br>units/mL | 300 units/mL | 1 |
| Aap Cas12b | 2 mg/mL or<br>15.4 uM | 500 nM | 1.625 |
| Aap Cas12b<br>crRNA | 360 ng/uL or<br>10 uM | 500 nM | 2.5 |
| Lateral Flow<br>Reporter | 100 uM | 125 nM | 0.0625 |
| Taurine | 500 mM | 50 mM | 5 |
| LAMP<br>primer mix | 10 X | 1 X | 5 |
| ddH<sub>2</sub>O |  |  | 11.8125 |
| **Total** |  |  | **45** |

\*\*\* NOTE: If lysing samples at 22 °C or 60 °C instead of 95 °C, replace 2 uL of ddH<sub>2</sub>O with 2 uL of Proteinase K Inhibitor working solution.
\*\*\* NOTE: If performing fluorescent readout, use the fluorescent reporter instead of the lateral flow reporter, at a final concentration of 250 nM.

For Step (3) reading out using lateral flow dipstick:

- HybriDetect Dipstick ([MGHD 1](https://www.milenia-biotec.com/en/product/hybridetect/)). Milenia Biotec GmbH

Positive control sequences

- SARS-CoV-2 RNA control (102019). Twist Bioscience

### Equipment

- 95 °C heat block or water bath
- 60 °C heat block or water bath
- Alternative: a sous vide immersion cooker capable of supporting the temperature range of 55 °C to 95 °C can also be used ([example](https://www.twistbioscience.com/coronavirus-research-tools)).

## One-step SHERLOCK Protocol for SARS-CoV-2 Detection

*\*\*\* IMPORTANT NOTE: To prevent sample contamination from confounding detection results, two different work areas should be used for performing Steps (1)/(2) and (3). Steps (1)/(2) should be performed in a pre-amplification area and is especially sensitive to contamination. Amplified samples should not be opened in the work area for Steps (1)/(2). A separate area for post-amplification reactions should be used for performing Step (3) of the protocol*.

*Step (1) – Lysis of patients sample*. *PERFORMED IN THE **<u>PRE</u>**-AMPLIFICATION AREA*

> Saliva or NP swab samples should be lysed using the QuickExtract lysis buffer.
>
> Mix 10 μL of saliva or NP swab sample with 10 μL of Quick Extract in an eppendorf tube.
>
> Incubate the sample-QuickExtract mixture at 95 °C for 5 minutes (or at room temperature or 60 °C for 10 mins) and proceed to Step (2).

*Step (2) – STOPCovid detection*. *PERFORMED IN THE **<u>PRE</u>**-AMPLIFICATION AREA*

> For each sample, set up one reaction as follows. In addition, a positive control can be set up using the SARS-CoV-2 control RNA. A negative control with Isothermal Amplification Buffer, MgSO_4_, dNTPs, Lateral Flow Reporter, and sample could also be set up to control for DNAse contamination that may produce false positive results. We did not observe DNAse contamination in negative control reactions when validating our assay using patient NP swabs.
>
> 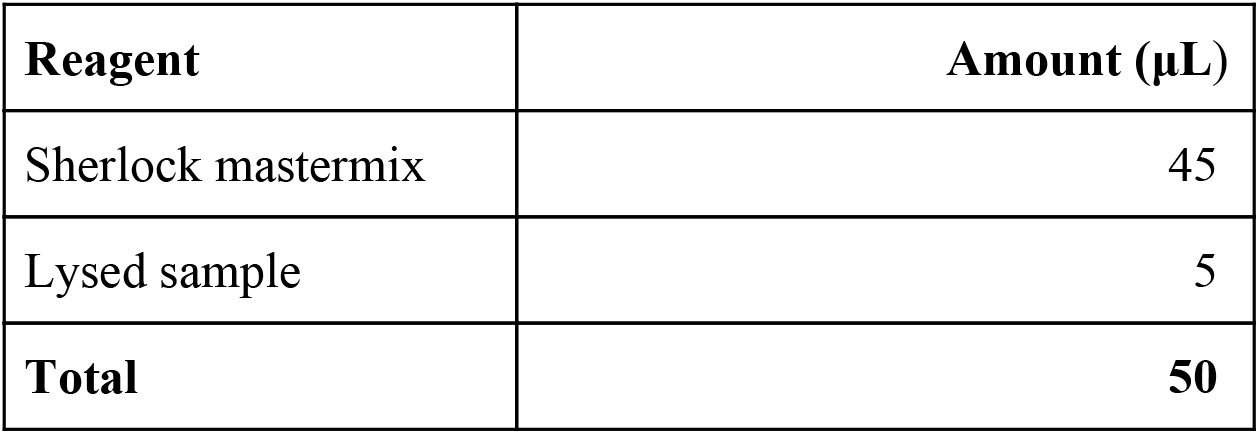
>
> Mix thoroughly and incubate each reaction at 60 °C for 1 hour. **Transfer the reaction tubes to the post-amplification area before proceeding to Step (3)**.

*Step (3) – Visual readout of detection result via lateral flow strip*. *PERFORMED IN THE **<u>POST</u>**-AMPLIFICATION AREA*

> Place a HybriDetect Dipstick into each reaction tube and wait for the reaction to flow through the dipstick. Generally, 2 minutes is enough time to develop a positive band at the limit of detection (longer wait times will not impact the result).
>
> Positive control samples should show the top line and a faint bottom line. Negative control samples should show the bottom line.
>
> For each test sample, check to see the top line appears, indicating positive SARS-CoV-2 detection.

## Additional Information

The protocol describes the use of lateral flow dipsticks for achieving a visual readout of the test result. For higher-throughput processing and to achieve semi-quantitative measurement of the virus load, the STOPCovid reaction can be performed with fluorescent readout by exchanging the reporter DNA. This modification allows measurement of hundreds of samples on a standard fluorescent plate reader. All of the reaction conditions remain the same except the lateral flow reporter should be replaced with a fluorescence reporter.

For additional information regarding SHERLOCK, a detailed general protocol can be found in the following reference:

- *[SHERLOCK: nucleic acid detection with CRISPR nucleases](https://www.nature.com/articles/s41596-019-0210-2)*. Kellner MJ, Koob JG, Gootenberg JS, Abudayyeh OO, and Zhang F. *Nature Protocols*. 2019 Oct;14(10):2986-3012. doi: 10.1038/s41596-019-0210-2.

## Discussion

STOPCovid is capable of rapid, point-of-care diagnosis of viral shedding in persons with COVID-19 disease. Our method achieved a LOD of 100 molecules of synthetic SARS-CoV-2 genomes in saliva or nasopharyngeal swab per reaction. Validation on patient samples showed that our method successfully diagnosed 12 positive and 5 negative COVID-19 patients, with at least 2 of 3 replicates scoring positive in infected persons.

Importantly, this simplified format, which has minimal sample processing steps and does not require complex instrumentation, can be performed by lay users, making it deployable in low-resource clinics, pharmacies, workplaces, and even developed for at-home use. For these types of applications, we anticipate that saliva samples, which have similar viral loads to nasopharyngeal swabs (Wyllie et al., 2020), will also work as input, as we have shown that spiked-in SARS-CoV-2 genomic RNA standards in healthy saliva is detected by STOPCovid.

Future development of a low-cost and modular heater capable of accommodating a disposable single-use test cartridge would further streamline the workflow and simplify the test for use in diverse environments such as at hospitals, nursing facilities, mobile testing stations, offices, and homes where the tests are most needed (Figure 6). The molecular components for STOPCovid are readily available and can be scaled with numerous commercial partners. The disposable test cartridge can be designed to be compatible with scalable manufacturing processes such as injection molding to allow for high throughput manufacturing. The cartridge would reduce both test complexity and risk of amplicon contamination by processing the sample input internally and directing the completed STOPCovid reaction mixture toward the lateral flow strip for readout. A mobile phone application can analyze images captured by the phone camera to readout test results (Figure 6).

**Figure 6:**
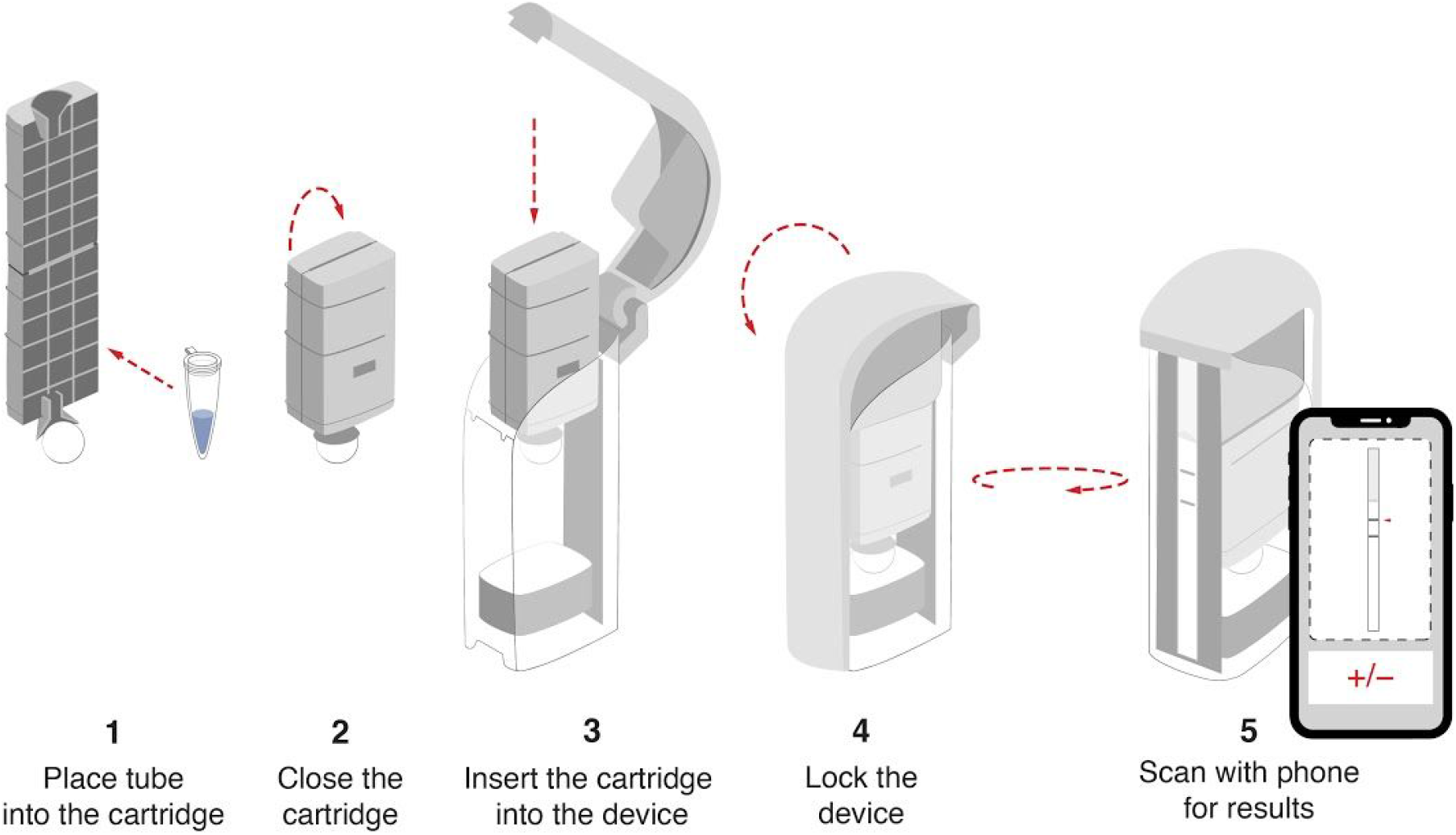
Device cartridge for reading out the STOPCovid test. In order to limit amplicon spread and contamination, sealed reaction tubes are placed in detection cartridges (Ustar, http://www.bioustar.com/en/about.aspx). User closes the cartridge and pulls the handle to release reaction onto a lateral flow strip. A mobile phone application can be used for quantitating the band intensity and determining the test result.

STOPCovid represents a promising platform for developing POC COVID-19 diagnostics and has the potential to play an important role in effective test-trace-isolate measures to end the COVID-19 pandemic and reopen society.

## Data Availability

Data is available at STOPCovid.science

http://www.STOPCovid.science

## Acknowledgements

We thank R. Shan of Quintara for oligo synthesis; K.Holden, A. Kadina, and J.Walker of Synthego for guide RNA synthesis; Abderrahim Farina for assistance with protein purification; and A. Tang for illustration assistance. Support for specimen collection was provided by Mark and Lisa Schwartz and Enid Schwartz. O.O.A, J.S.G., and F.Z. are supported by the Patrick J. McGovern Foundation and DARPA Award D18AC00006. F.Z is also supported by the NIH (1R01-MH110049 and 1DP1-HL141201 grants); Mathers Foundation; the Howard Hughes Medical Institute; Open Philanthropy Project; J. and P. Poitras; and R. Metcalfe.

## Ethical Statement

This study was performed under the MIT IRB IRB-6095, the University of Washington IRB STUDY00010205, and Partners Healthcare IRB #2020P000804.

